# A phase 2, open-label, single-arm monotherapy trial of Sulfasalazine in patients with PancrEatic AdenocaRcinoma: Statistical analysis plan for the SPEAR study

**DOI:** 10.64898/2025.12.04.25341652

**Authors:** Stacey Llewellyn, Chris Gianacas, Frank Lin, David Goldstein

## Abstract

**Background:** Pancreatic ductal adenocarcinoma (PDAC) is associated with an extremely poor prognosis, with a 5-year survival rate of less than 9% (1). The SPEAR study is a Phase II, single-arm, open-label, signal-seeking trial evaluating the clinical activity of sulfasalazine, a repurposed anti-inflammatory agent, in patients with advanced or metastatic PDAC who have progressed on one prior line of systemic therapy. This document extends the published study protocol by pre-specifying the planned statistical analyses, including post hoc analyses following early trial termination for futility.

**Design and Setting:** Single-arm, multi-centre Phase II trial conducted in two sequential cohorts (Cohort 1: n=17; Cohort 2: n=17–22), with inclusion contingent on remaining on study until at least Cycle 2 Day 1 (C2D1). Sulfasalazine was administered using a pharmacokinetically guided dose-escalation strategy, with escalation speed informed by acetylator status to minimise toxicity. Interim analyses were planned to assess early signals of efficacy and safety, with predefined stopping rules.

**Outcomes:** The primary outcome is the proportion of patients achieving progression-free survival (PFS) at 6 months. Secondary outcomes include objective tumour response rate, median PFS, overall survival (OS) at 12 months, median OS, time to progression, safety, and dose intensity. Tertiary outcomes include exploratory biomarkers such as tumour markers (CA 19-9, CEA), peripheral blood glutathione, and serum collagen degradation products.

**Planned Analyses:** All analyses will be conducted using R (version 4.0.0 or above) or SAS Enterprise Guide (version 8.3 or above), with a two-sided alpha of 0.05. Binary outcomes, including the primary outcome, will be summarised as proportions with 95% confidence intervals. Time-to-event outcomes will be analysed using Kaplan-Meier methods, and continuous outcomes will be summarised using appropriate descriptive statistics. Post hoc exploratory analyses will examine factors associated with recruitment, clinical suitability, and early treatment discontinuation. These include evaluating the relationship between line of therapy and time on study and assessing whether baseline HRQoL or inflammatory biomarkers may better inform future eligibility criteria. Additional analyses will explore associations between baseline characteristics and overall survival or early discontinuation using regression and survival methods. All post hoc analyses will be clearly identified and interpreted as hypothesis-generating.

## 2 Study background

The full SPEAR study is described in detail in the study protocol. Briefly, the SPEAR study is a phase II, single-arm, open-label monotherapy trial to investigate the clinical activity of sulfasalazine in patients with pancreatic ductal adenocarcinoma (PDAC), using a signal-seeking design. Sulfasalazine is a well-established drug that has been repurposed as an innovative approach to treat PDAC, specifically in patients with advanced or metastatic pancreatic cancer that have progressed on one-line of prior systemic therapy. The aim of the study is to assess the clinical activity of sulfasalazine treatment in patients with PDAC who remain on study until at least Cycle 2 Day 1 (C2D1), as measured by PFS at 6 months as a primary objective and objective response as a secondary objective. Participants will be followed up until death or trial closure (whichever occurs first).

The SPEAR study will adopt a pharmacokinetic approach to initial dosing of sulfasalazine. Once tolerance of 1.5g per day is established, the dose is incrementally increased over several days until reaching the target dose (4.5g per day in three divided doses for participants ≥50kg), subject to tolerability. Acetylator status (slow or fast) will be used to inform the speed of sulfasalazine dose escalation. To minimise toxicity, dose escalation will be slower in slow acetylators.

### 2.1 Sample size and Statistical Significance Boundary

We assumed a null hypothesis of PFS at 6 months rate equal to 10% (based on the 5 FU/LV arms of NAPOLI1 trial (2)) to be tested against a one-sided alternative hypothesis that the PFS rate is 26% with 80% power and 5% type-I error rate. A Fleming 3-stage single-arm trial design will be used to test whether the proportion responding (P) warrants continuation to the next phase (H0: P ≤ 0.1 versus H1: P ≥ 0.26) (3, 4).

Planned recruitment for the SPEAR trial consisted of two cohorts, Cohort 1 (n=17) and Cohort 2 (n=17-22), recruited successively, with participant inclusion in the trial provided C2D1 visit completion. Following a signal-seeking study design, one preliminary data review and two interim analyses are planned to provide an early signal of clinical anti-tumour activity and safety of sulfasalazine, as follows:

- Preliminary review: Cohort 1A (n=13), all C2D1 data available

`o` Stopping criteria

▪ Futility: the number of participants with PFS at 2 months is equal to zero
- Interim analysis one: cohort 1A (n=13), all primary end point data available (6-month, PFS (C6D28))

`o` Stopping criteria

▪ Futility: the number of participants with PFS at 6 months is equal to zero
▪ Efficacy: the number of participants with PFS at 6 months ≥5
`o` Recruitment not paused. Continued recruitment to Cohort 1B.
- Interim analysis two: cohort 1A/B (n=17), all primary end point data available (6-month, PFS (C6))

`o` Stopping criteria

▪ Futility: the number of participants with PFS at 6 months ≤1in Cohort 1B (n=4)
▪ Efficacy: the number of participants with PFS at 6 months ≥6 in Cohort 1A/B (n=17)
`o` Recruitment to Cohort 2 paused until interim analysis two results are available.

## 3 Study aims and objectives

### Aim

The overall aim of the study is to investigate the clinical activity of sulfasalazine in patients with PDAC using a signal-seeking trial design.

### Primary Objective

To quantify PFS rate at 6 months among patients with PDAC treated with sulfasalazine.

### Secondary Objectives

Among patients with PDAC treated with sulfasalazine to quantify:

1. Objective tumour response rate
2. Median PFS
3. Clinical benefit rate (CBR) at 16 weeks *
4. Duration of clinical benefit (DCB) *
5. OS rate at 12 months
6. Median OS
7. Time to progression (TTP)
8. Median Growth Modulation Index (GMI) *
9. Safety of treatment
10. Tolerability of treatment *
11. HRQOL over the course of the trial *
12. Dose intensity achieved (mean, range and percentage of intended/target dose)
13. Median duration of objective tumour response *

### Tertiary Objectives

Among patients with PDAC treated with sulfasalazine to quantify:

1. Tumour fibrosis *
2. Intratumoural glutathione *
3. Tumour marker responses (CA 19-9 or Carcinoembryonic antigen [CEA] in patients that are non-secretors of CA 19-9) and their associations with clinical and radiological endpoints.
4. Peripheral blood glutathione
5. Serum collagen degradation products
6. Association of peripheral blood glutathione and serum collagen degradation products
7. Stromal and/or tumour SLC7A11 *
8. Tumour metabolic response using fluorodeoxyglucose-positron emission tomography (FDG-PET) *

* Out of scope for this statistical analysis plan.

Note: Some additional exploratory objectives were noted in the protocol however these are out of scope for this SAP. Similarly, while tertiary endpoints will be presented descriptively, association of tertiary endpoints with clinical endpoints are out of scope for this SAP.

## 4 Study endpoints

The clinical details for each endpoint can be found in protocol section five.

### 4.1 Primary endpoint

The primary outcome, progression free survival (PFS) at 6 months, is defined as the proportion of participants who are alive and progression free at the end of cycle six (approximately day 168 from start of study treatment). Progression is defined as first evidence of disease progression (clinical or radiological) or death from any cause, whichever occurs first (full details available in the protocol section 5.1.1).

### 4.2 Secondary outcomes

#### 4.2.1 Objective Tumour Response Rate

The objective tumour response rate is defined as the proportion of participants who achieved an objective response at any time point (see protocol section 5.2.1 for clinical definitions).

#### 4.2.2 Median Progression Free Survival

Median PFS is similar to the primary outcome but as a time-to-event endpoint. Participants who did not progress or die will be censored on the date of their last clinical assessment or tumour assessment.

#### 4.2.3 Overall Survival Rate at 12 Months

Overall Survival (OS) rate at 12 months is defined as the proportion of participants who are alive at 12 months from start date of study treatment to date of death from any cause, or the date of last known vital status during follow up within 12 months from start date of study treatment.

#### 4.2.4 Median Overall Survival

OS is defined as the interval from the start date of study treatment to date of death from any cause. Participants who did not die will be censored at the date of last known follow-up alive. Like median PFS, this is a time-to-event outcome.

#### 4.2.5 Median Time to Progression

Median Time to Progression (TTP) is defined as the time interval from the start date of study treatment to the date of first evidence of disease progression on the SPEAR trial or death due to cancer. Like median PFS, this is a time-to-event outcome.

#### 4.2.6 Safety of Treatment

The safety of sulfasalazine treatment will be assessed by the incidence and rates of AEs and serious AEs (SAEs).

An AE is deemed to be due to treatment if the causal relationship between the event and sulfasalazine treatment is judged by the Investigator to be definitely, probably or possibly related.

#### 4.2.7 Dose Intensity Achieved

Dose intensity will be evaluated as a continuous endpoint, defined as the absolute drug dose delivered per unit of time and expressed in grams per week. Dose intensity at each timepoint will be presented. In addition, the total dose administered will be reported as a proportion of the target dose, both at each timepoint and across the overall study duration

### 4.3 Tertiary outcomes

Potential biomarkers that may correlate with the clinical activity of sulfasalazine will be evaluated as tertiary outcomes measures (see protocol section 5.3 for clinical details)

#### 4.3.1 Peripheral Blood Glutathione

These are continuous values that will be calculated pre- and post-treatment.

#### 4.3.2 Tumour Marker CA 19-9

This is a continuous value that will be measured at baseline, on day 1 of each treatment cycle, and at disease progression

#### 4.3.3 Tumour Marker CEA

This is a continuous value that will be measured for CA 19-9 negative patients at baseline and, for a subset of those patients, on day 1 of each treatment cycle, and at disease progression (see section 5.3.4 of the protocol for full details).

#### 4.3.4 Serum Collagen Degradation Products

This is a continuous value that will be calculated pre- and post-treatment.

## 5 Analyses

All analyses will be conducted using R (version 4.0.0 or above) or SAS Enterprise Guide (version 8.3 or above). The significance level (alpha) will be set to 0.05 (two sided).

### 5.1 Populations for Analysis

#### 5.1.1 Screened Set

The *Screened Set* includes all patients who provided informed consent and were assigned a screening number, regardless of eligibility or treatment status.

#### 5.1.2 Consented Set

The *Consented Set* includes all patients who met eligibility criteria and were formally enrolled in the study, regardless of whether they received study treatment.

#### 5.1.3 Safety Set

The *Safety Set* consists of all patients who received at least one dose of study treatment.

#### 5.1.4 Primary Analysis Set

The *Primary Analysis Set* is the main efficacy population, defined as all patients in the Consented Set who reached cycle 2 day 1 (C2D1) of the treatment protocol.

### 5.2 Subject disposition

The flow of patients through the trial will be displayed in a CONSORT diagram (5). The report will include the number of participants who: were assessed for eligibility (including reason for exclusion), were enrolled, were administered the intervention at cycle 1 day 1 (C1D1), reached cycle 2 day 1 (C2D1) for inclusion in the study, and who reached the primary outcome timepoint of cycle 6 day 28 (C6D28). Details of interim analysis one and early stopping criteria are also included. The number of participants with disease progression or died, lost to follow-up/ withdrew from the study as well as the reason for withdrawal will be provided at each of these identified timepoints.

### 5.3 Baseline characteristics

Participant baseline characteristics will be presented overall for the *Safety Set, Consented Set* and the *Primary Analysis Set*. Categorical variables will be presented as frequency and percentages, with percentages calculated according to the number of participants for who data are available. Continuous variables will be presented as mean (standard deviation (SD)), and median and interquartile range (Q1-Q3). The following baseline measures will be included in summaries: sex, age (years), ethnic group, height (cm), weight (kg), BMI (kg/m2), smoking status and regularity, acetylator status, important comorbidities (e.g. diabetes), and Eastern Cooperative Oncology Group (ECOG) performance status.

Participants’ history of pancreatic cancer will similarly be presented overall. The following measures will be included in summaries: primary site histology, current extent of disease, HRQOL summaries, summary of prior CA 19-9 and CEA levels pre-first line treatment, at nadir, and at cessation of first-line treatment, and summaries of previous cancer treatments, including indication, reason for stopping and number of cycles completed for systemic therapy.

### 5.4 Primary endpoint

As this is a single-arm study, the primary endpoint will be reported as counts and proportions (percentage and 95% confidence intervals (6)) of patients who did or did not achieve PFS at six months in the *Primary Analysis Set*.

### 5.5 Secondary endpoints

All secondary endpoints will be analysed using the *Primary Analysis Set*.

Binary secondary outcomes will be estimated as described in the primary analysis. Binary outcomes in this study include objective tumour response rate, and OS rate at 12 months.

Time to event outcomes, will be summarised using the Kaplan-Meier method (7). Time to event outcomes include median PFS, median OS, and TTP. Results will be reported as the median (95% CI), accompanied by the number of observed events and number of censored observations.

Continuous outcomes will be summarised by the median, range, 25th/75th percentiles and mean (with standard deviation) at required timepoints. Continuous outcomes include total dose administered, and dose intensity achieved.

### 5.6 Tertiary endpoints

Generally, tertiary endpoints will be calculated as per the secondary endpoints using the *Primary Analysis Set*. Endpoints available at multiple time points will be provided as figures with 95% confidence intervals. Endpoints available before and after treatment will be presented as tables.

### 5.7 Post hoc Analyses

Due to early termination of the trial at the interim analysis for futility, additional exploratory (post hoc) analyses will be conducted to support interpretation of the study findings. These analyses were not pre-specified in the protocol and are intended to explore factors influencing recruitment and clinical suitability. All post hoc analyses will be clearly identified as such and interpreted accordingly (8).

To support this, an exploratory analysis will assess the relationship between line of therapy at enrolment and time on study using the *Consented Set*. Specifically, time on study will be compared between patients who were truly second-line (single systemic therapy), and those who were beyond second-line (two or three systemic therapies) but had good performance status (ECOG 0-1). Time on study will be defined as the interval from the date of enrolment to the date of patient discontinuation from the study or death due to any cause.

Time on study, as a continuous outcome, will be analysed using linear regression with line of therapy as a predictor. Given the exploratory nature and small sample size, the model will not adjust for additional covariates. Model assumptions including linearity, homoscedasticity and normality of residuals will be assessed, and if violated, appropriate transformations or non-parametric methods will be considered. Results will be reported as the estimated mean difference in time on study between groups, along with 95% confidence intervals and p-values.

If differences in time on study are observed between patients who were truly second-line and those who were beyond second-line (despite ECOG of 0 or 1), additional exploratory analysis may be conducted to examine alternative or complementary indicators of clinical suitability for trial inclusion. These analyses will aim to:

- Evaluate whether baseline HRQoL, using the QLQ-C30 Summary score, is more strongly associated with time on study than ECOG performance status.
- Investigate whether baseline biomarkers, including albumin, white cell count, or the derived neutrophil-to-lymphocyte ratio (NLR), may provide better prognostic value or inform future eligibility criteria.

For each variable, the association with time on study will be assessed using univariate linear regression. If model assumptions are not met, as described above, Spearman correlation will be used instead. Estimates, 95% confidence intervals, and p-values will be presented.

Analysis will also be conducted using the *Consented Set* to explore associations between overall survival and baseline characteristics: tumour metabolic response, SLC7A11, and serum collagen degradation. Overall survival will be summarised using Kaplan–Meier methods, with comparisons between dichotomised baseline groups made using log-rank or, if appropriate, alternative weighted log-rank tests (e.g. Peto–Peto) based on the observed data (7). Results will include Kaplan-Meier curves, median survival times with 95% confidence intervals, number of events and at-risk patients at each time point, and log-rank/weighted log-rank test results (including chi-square statistics and p-value).

The association between patient baseline characteristics and the likelihood of remaining on therapy for more than one month (binary endpoint: ≤1 month vs. >1 month) will be assessed using logistic regression. This analysis will be conducted on the *Safety Set*, defined as all patients who received at least one dose of study treatment. Baseline characteristics considered as potential predictors include platelet-to-lymphocyte ratio (POR), white cell count (WCC), serum albumin, and liver function test results. Variables will be evaluated in univariable logistic regression models, with odds ratios (ORs) with 95% confidence intervals (CIs) reported. Model fit will be assessed using the Hosmer–Lemeshow goodness-of-fit test, and predictive discrimination will be evaluated using the area under the receiver operating characteristic curve (AUC) (9).

These analyses are hypothesis-generating and intended to inform future study design. Given the small sample size within each respective analysis set, all findings will be interpreted with caution and treated as exploratory. Findings will be transparently reported in the final study report and any resulting publications.

### 5.8 Safety outcomes

The Safety of Treatment outcomes of AEs and serious AEs (SAEs) will be summarised using the *Safety Set*, defined as all patients who received at least one dose of study treatment. The number and proportion of patients experiencing at least one event will be reported, as well as the total number of events experienced.

Additional classifications of AEs will be similarly summarized, including causal relationship to study medication, severity (NCI CTCAE v 5.0 graded), expected AEs, event status, action taken, dose down-titration and reaction abatement and reappearance following treatment adjustments.

A listing of all AEs and SAEs will be reported (in an appendix).

## Data Availability

This manuscript describes the planned statistical analyses; no study data are included or available at this time.

## 7 Proposed Tables and Figures

**Figure 1.**
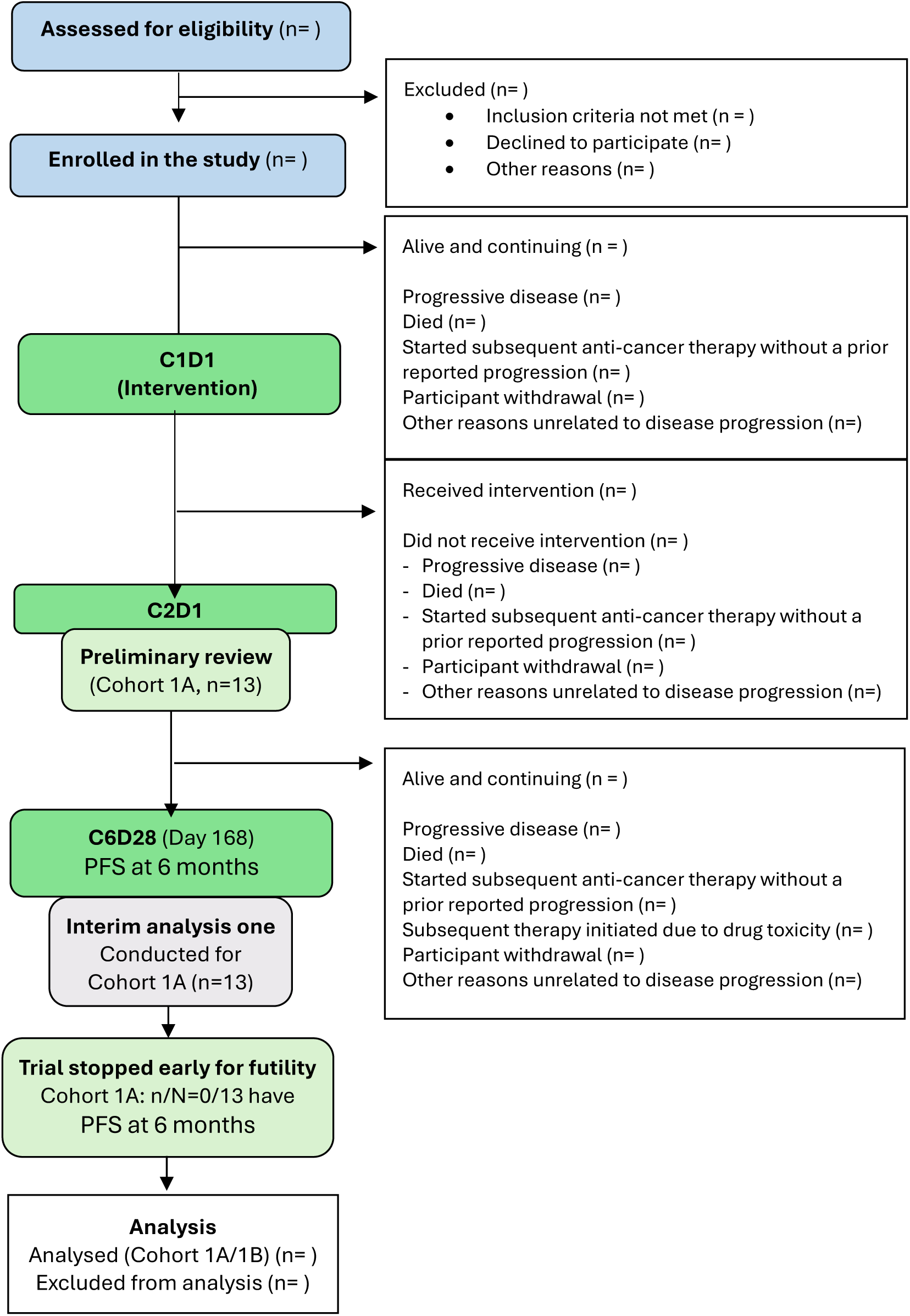
Consort flow diagram

**Figure 2.** Kaplan-Meier plot of progression free survival (PFS) in the Primary Analysis Set

**Figure 3.** Kaplan-Meier plot of overall survival (OS) in the Primary Analysis Set

**Figure 4.** Kaplan-Meier plot of time to tumour progression in the Primary Analysis Set

**Figure 5.** Swimmer plot of patient progression following enrolment in the Safety Set

**Figure 6.** Kaplan-Meier curves for overall survival stratified by baseline tumour metabolic response in the Consented Set [post-hoc analysis]

**Figure 7.** Kaplan-Meier curves for overall survival stratified by baseline SLC7A11 status in the Consented Set [post-hoc analysis]

**Figure 8.** Kaplan-Meier curves for overall survival stratified by baseline serum collagen degradation marker levels in the Consented Set [post-hoc analysis]

**Table 1.**
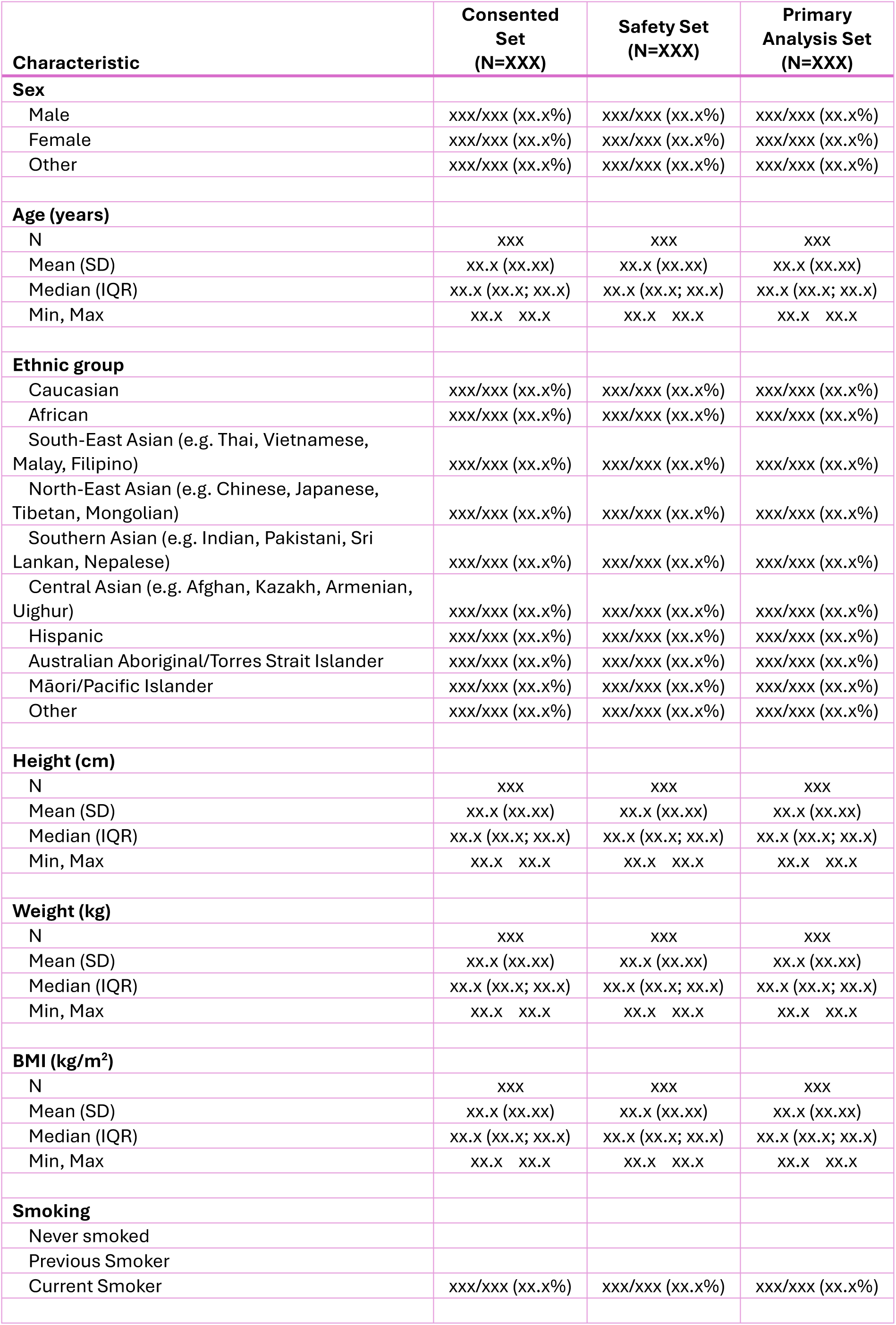
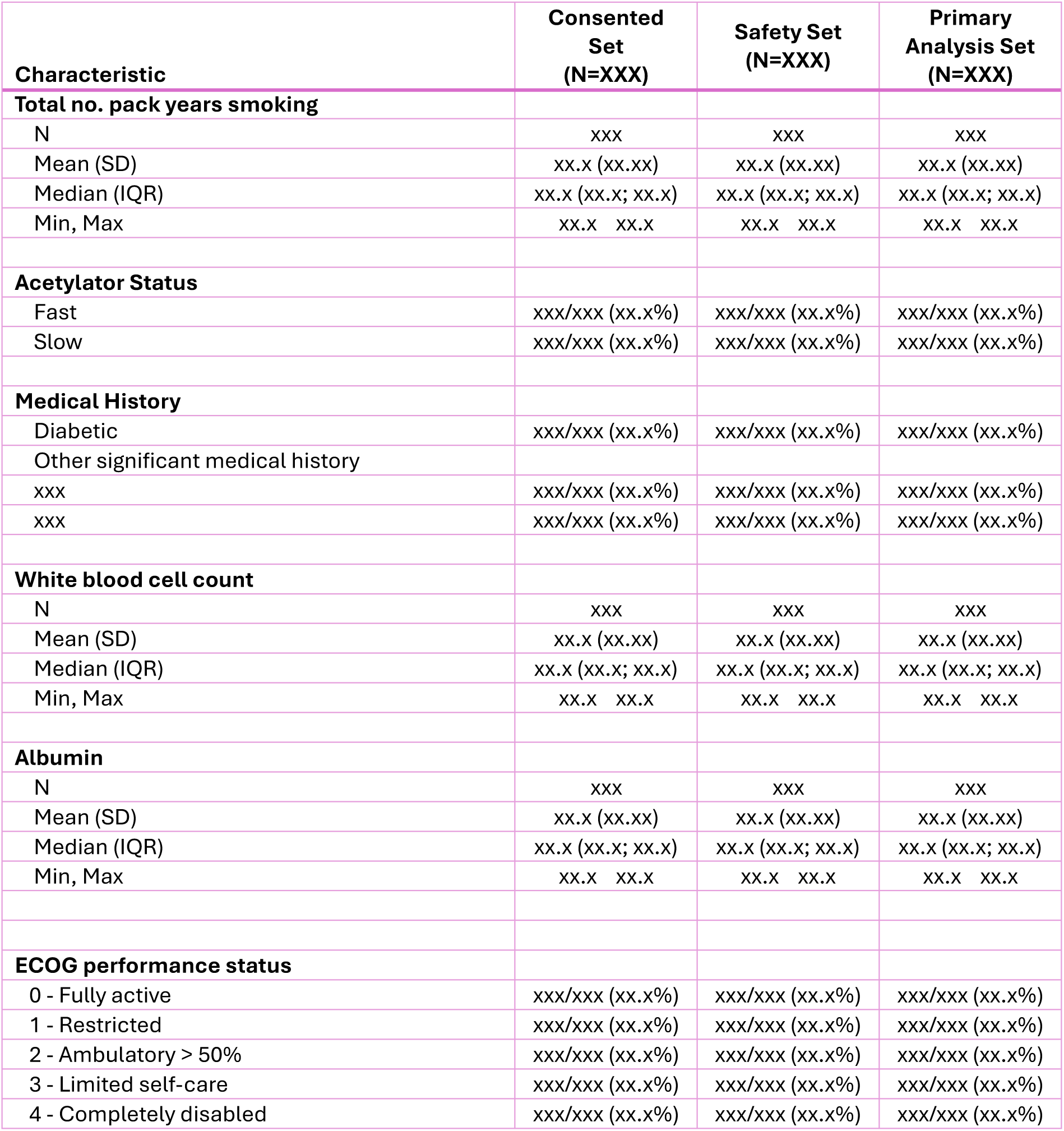
Baseline characteristics.

| Characteristic | Consented Set<br>(N=XXX) | Safety Set<br>(N=XXX) | Primary Analysis Set<br>(N=XXX) |
| --- | --- | --- | --- |
| **Sex** |  |  |  |
| Male | xxx/xxx (xx.x%) | xxx/xxx (xx.x%) | xxx/xxx (xx.x%) |
| Female | xxx/xxx (xx.x%) | xxx/xxx (xx.x%) | xxx/xxx (xx.x%) |
| Other | xxx/xxx (xx.x%) | xxx/xxx (xx.x%) | xxx/xxx (xx.x%) |
| **Age (years)** |  |  |  |
| N | xxx | xxx | xxx |
| Mean (SD) | xx.x (xx.xx) | xx.x (xx.xx) | xx.x (xx.xx) |
| Median (IQR) | xx.x (xx.x; xx.x) | xx.x (xx.x; xx.x) | xx.x (xx.x; xx.x) |
| Min, Max | xx.x xx.x | xx.x xx.x | xx.x xx.x |
| **Ethnic group** |  |  |  |
| Caucasian | xxx/xxx (xx.x%) | xxx/xxx (xx.x%) | xxx/xxx (xx.x%) |
| African | xxx/xxx (xx.x%) | xxx/xxx (xx.x%) | xxx/xxx (xx.x%) |
| South-East Asian (e.g. Thai, Vietnamese, Malay, Filipino) | xxx/xxx (xx.x%) | xxx/xxx (xx.x%) | xxx/xxx (xx.x%) |
| North-East Asian (e.g. Chinese, Japanese, Tibetan, Mongolian) | xxx/xxx (xx.x%) | xxx/xxx (xx.x%) | xxx/xxx (xx.x%) |
| Southern Asian (e.g. Indian, Pakistani, Sri Lankan, Nepalese) | xxx/xxx (xx.x%) | xxx/xxx (xx.x%) | xxx/xxx (xx.x%) |
| Central Asian (e.g. Afghan, Kazakh, Armenian, Uighur) | xxx/xxx (xx.x%) | xxx/xxx (xx.x%) | xxx/xxx (xx.x%) |
| Hispanic | xxx/xxx (xx.x%) | xxx/xxx (xx.x%) | xxx/xxx (xx.x%) |
| Australian Aboriginal/Torres Strait Islander | xxx/xxx (xx.x%) | xxx/xxx (xx.x%) | xxx/xxx (xx.x%) |
| Māori/Pacific Islander | xxx/xxx (xx.x%) | xxx/xxx (xx.x%) | xxx/xxx (xx.x%) |
| Other | xxx/xxx (xx.x%) | xxx/xxx (xx.x%) | xxx/xxx (xx.x%) |
| **Height (cm)** |  |  |  |
| N | xxx | xxx | xxx |
| Mean (SD) | xx.x (xx.xx) | xx.x (xx.xx) | xx.x (xx.xx) |
| Median (IQR) | xx.x (xx.x; xx.x) | xx.x (xx.x; xx.x) | xx.x (xx.x; xx.x) |
| Min, Max | xx.x xx.x | xx.x xx.x | xx.x xx.x |
| **Weight (kg)** |  |  |  |
| N | xxx | xxx | xxx |
| Mean (SD) | xx.x (xx.xx) | xx.x (xx.xx) | xx.x (xx.xx) |
| Median (IQR) | xx.x (xx.x; xx.x) | xx.x (xx.x; xx.x) | xx.x (xx.x; xx.x) |
| Min, Max | xx.x xx.x | xx.x xx.x | xx.x xx.x |
| **BMI (kg/m<sup>2</sup>)** |  |  |  |
| N | xxx | xxx | xxx |
| Mean (SD) | xx.x (xx.xx) | xx.x (xx.xx) | xx.x (xx.xx) |
| Median (IQR) | xx.x (xx.x; xx.x) | xx.x (xx.x; xx.x) | xx.x (xx.x; xx.x) |
| Min, Max | xx.x xx.x | xx.x xx.x | xx.x xx.x |
| **Smoking** |  |  |  |
| Never smoked |  |  |  |
| Previous Smoker |  |  |  |
| Current Smoker | xxx/xxx (xx.x%) | xxx/xxx (xx.x%) | xxx/xxx (xx.x%) |
| **Total no. pack years smoking** |  |  |  |
| N | xxx | xxx | xxx |
| Mean (SD) | xx.x (xx.xx) | xx.x (xx.xx) | xx.x (xx.xx) |
| Median (IQR) | xx.x (xx.x; xx.x) | xx.x (xx.x; xx.x) | xx.x (xx.x; xx.x) |
| Min, Max | xx.x xx.x | xx.x xx.x | xx.x xx.x |
| **Acetylator Status** |  |  |  |
| Fast | xxx/xxx (xx.x%) | xxx/xxx (xx.x%) | xxx/xxx (xx.x%) |
| Slow | xxx/xxx (xx.x%) | xxx/xxx (xx.x%) | xxx/xxx (xx.x%) |
| **Medical History** |  |  |  |
| Diabetic | xxx/xxx (xx.x%) | xxx/xxx (xx.x%) | xxx/xxx (xx.x%) |
| Other significant medical history |  |  |  |
| xxx | xxx/xxx (xx.x%) | xxx/xxx (xx.x%) | xxx/xxx (xx.x%) |
| xxx | xxx/xxx (xx.x%) | xxx/xxx (xx.x%) | xxx/xxx (xx.x%) |
| **White blood cell count** |  |  |  |
| N | xxx | xxx | xxx |
| Mean (SD) | xx.x (xx.xx) | xx.x (xx.xx) | xx.x (xx.xx) |
| Median (IQR) | xx.x (xx.x; xx.x) | xx.x (xx.x; xx.x) | xx.x (xx.x; xx.x) |
| Min, Max | xx.x xx.x | xx.x xx.x | xx.x xx.x |
| **Albumin** |  |  |  |
| N | xxx | xxx | xxx |
| Mean (SD) | xx.x (xx.xx) | xx.x (xx.xx) | xx.x (xx.xx) |
| Median (IQR) | xx.x (xx.x; xx.x) | xx.x (xx.x; xx.x) | xx.x (xx.x; xx.x) |
| Min, Max | xx.x xx.x | xx.x xx.x | xx.x xx.x |
| **ECOG performance status** |  |  |  |
| 0 - Fully active | xxx/xxx (xx.x%) | xxx/xxx (xx.x%) | xxx/xxx (xx.x%) |
| 1 - Restricted | xxx/xxx (xx.x%) | xxx/xxx (xx.x%) | xxx/xxx (xx.x%) |
| 2 - Ambulatory > 50% | xxx/xxx (xx.x%) | xxx/xxx (xx.x%) | xxx/xxx (xx.x%) |
| 3 - Limited self-care | xxx/xxx (xx.x%) | xxx/xxx (xx.x%) | xxx/xxx (xx.x%) |
| 4 - Completely disabled | xxx/xxx (xx.x%) | xxx/xxx (xx.x%) | xxx/xxx (xx.x%) |

**Table 2.** Pancreatic cancer history.

| Clinical characteristic | Consented Set (N=XXX) | Safety Set (N=XXX) | Primary Analysis Set (N=XXX) |
| --- | --- | --- | --- |
| **Primary site histology** |  |  |  |
| Adenocarcinoma | xxx/xxx (xx.x%) | xxx/xxx (xx.x%) | xxx/xxx (xx.x%) |
| Mixed | xxx/xxx (xx.x%) | xxx/xxx (xx.x%) | xxx/xxx (xx.x%) |
| **Current Extent of Disease** |  |  |  |
| **Locally Advanced** | xxx/xxx (xx.x%) | xxx/xxx (xx.x%) | xxx/xxx (xx.x%) |
| **Regional lymph node involvement** | xxx/xxx (xx.x%) | xxx/xxx (xx.x%) | xxx/xxx (xx.x%) |
| **Distant metastases** | xxx/xxx (xx.x%) | xxx/xxx (xx.x%) | xxx/xxx (xx.x%) |
| **Liver metastases present** | xxx/xxx (xx.x%) | xxx/xxx (xx.x%) | xxx/xxx (xx.x%) |
| **Participant symptomatic** | xxx/xxx (xx.x%) | xxx/xxx (xx.x%) | xxx/xxx (xx.x%) |
| **Stromal &/or Tumour SLC7A11** |  |  |  |
| None | xxx/xxx (xx.x%) | xxx/xxx (xx.x%) | xxx/xxx (xx.x%) |
| Low | xxx/xxx (xx.x%) | xxx/xxx (xx.x%) | xxx/xxx (xx.x%) |
| Medium | xxx/xxx (xx.x%) | xxx/xxx (xx.x%) | xxx/xxx (xx.x%) |
| High | xxx/xxx (xx.x%) | xxx/xxx (xx.x%) | xxx/xxx (xx.x%) |
| **Tumour Metabolic Response** |  |  |  |
| Complete metabolic response | xxx/xxx (xx.x%) | xxx/xxx (xx.x%) | xxx/xxx (xx.x%) |
| Partial metabolic response | xxx/xxx (xx.x%) | xxx/xxx (xx.x%) | xxx/xxx (xx.x%) |
| Stable metabolic disease | xxx/xxx (xx.x%) | xxx/xxx (xx.x%) | xxx/xxx (xx.x%) |
| Progressive metabolic disease | xxx/xxx (xx.x%) | xxx/xxx (xx.x%) | xxx/xxx (xx.x%) |
| **Prior CA 19-9** |  |  |  |
| **Pre-first line treatment** |  |  |  |
| N | xxx | xxx | xxx |
| Mean (SD) | xx.x (xx.xx) | xx.x (xx.xx) | xx.x (xx.xx) |
| Median (IQR) | xx.x (xx.x; xx.x) | xx.x (xx.x; xx.x) | xx.x (xx.x; xx.x) |
| Min, Max | xx.x xx.x | xx.x xx.x | xx.x xx.x |
| **Nadir** |  |  |  |
| N | xxx | xxx | xxx |
| Mean (SD) | xx.x (xx.xx) | xx.x (xx.xx) | xx.x (xx.xx) |
| Median (IQR) | xx.x (xx.x; xx.x) | xx.x (xx.x; xx.x) | xx.x (xx.x; xx.x) |
| Min, Max | xx.x xx.x | xx.x xx.x | xx.x xx.x |
| **At cessation of first-line treatment** |  |  |  |
| N | xxx | xxx | xxx |
| Mean (SD) | xx.x (xx.xx) | xx.x (xx.xx) | xx.x (xx.xx) |
| Median (IQR) | xx.x (xx.x; xx.x) | xx.x (xx.x; xx.x) | xx.x (xx.x; xx.x) |
| Min, Max | xx.x xx.x | xx.x xx.x | xx.x xx.x |
| **Prior CEA** |  |  |  |
| **Pre-first line treatment** |  |  |  |
| N | xxx | xxx | xxx |
| Mean (SD) | xx.x (xx.xx) | xx.x (xx.xx) | xx.x (xx.xx) |

| Clinical characteristic | Consented Set<br>(N=XXX) | Safety Set<br>(N=XXX) | Primary Analysis Set<br>(N=XXX) |
| --- | --- | --- | --- |
| Median (IQR) | xx.x (xx.x; xx.x) | xx.x (xx.x; xx.x) | xx.x (xx.x; xx.x) |
| Min, Max | xx.x xx.x | xx.x xx.x | xx.x xx.x |
| **Nadir** |  |  |  |
| N | xxx | xxx | xxx |
| Mean (SD) | xx.x (xx.xx) | xx.x (xx.xx) | xx.x (xx.xx) |
| Median (IQR) | xx.x (xx.x; xx.x) | xx.x (xx.x; xx.x) | xx.x (xx.x; xx.x) |
| Min, Max | xx.x xx.x | xx.x xx.x | xx.x xx.x |
| **At cessation of first-line treatment** |  |  |  |
| N | xxx | xxx | xxx |
| Mean (SD) | xx.x (xx.xx) | xx.x (xx.xx) | xx.x (xx.xx) |
| Median (IQR) | xx.x (xx.x; xx.x) | xx.x (xx.x; xx.x) | xx.x (xx.x; xx.x) |
| Min, Max | xx.x xx.x | xx.x xx.x | xx.x xx.x |
| **Time from diagnosis to study enrolment (months)** |  |  |  |
| N | xxx | xxx | xxx |
| Mean (SD) | xx.x (xx.xx) | xx.x (xx.xx) | xx.x (xx.xx) |
| Median (IQR) | xx.x (xx.x; xx.x) | xx.x (xx.x; xx.x) | xx.x (xx.x; xx.x) |
| Min, Max | xx.x xx.x | xx.x xx.x | xx.x xx.x |

**Table 3.**
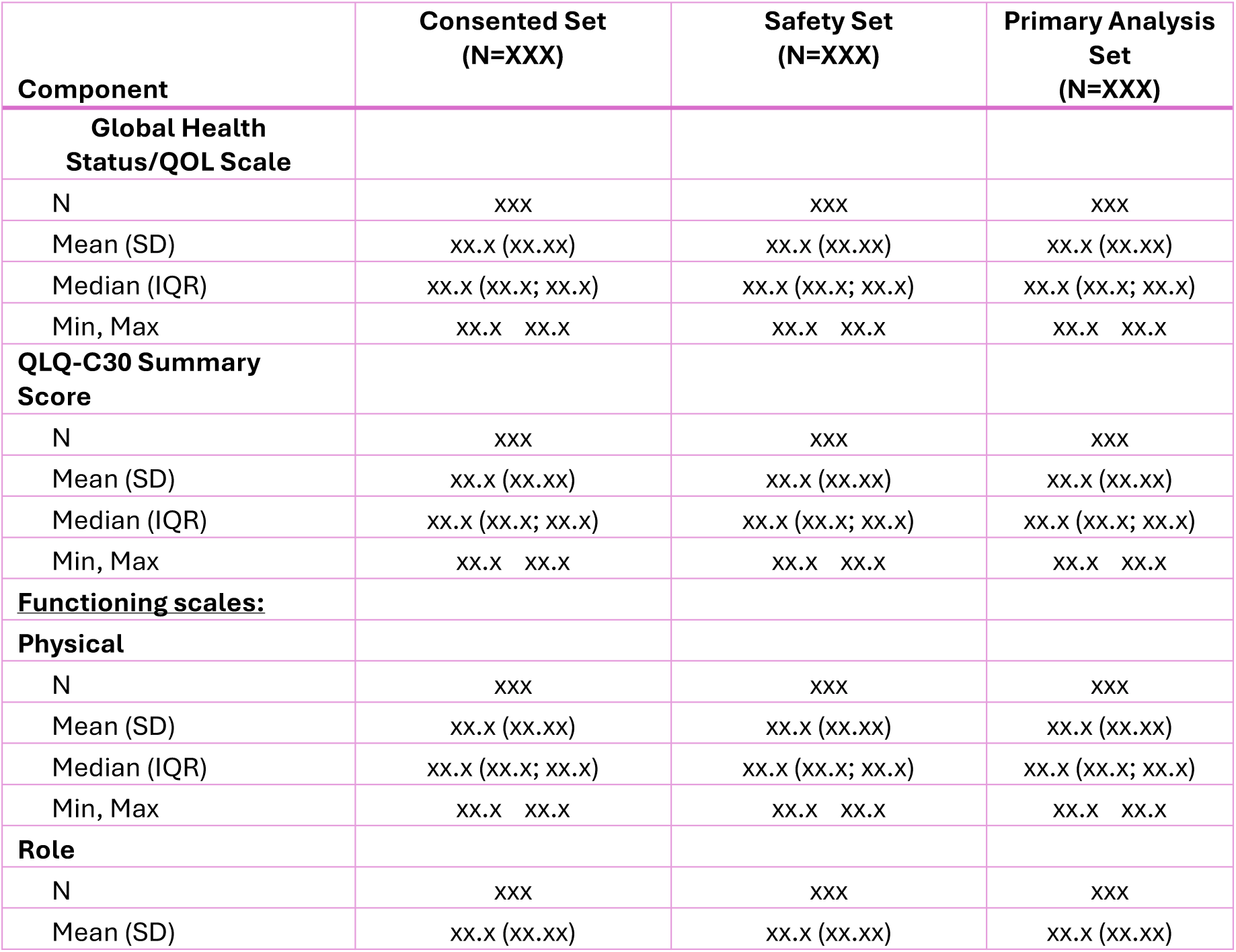
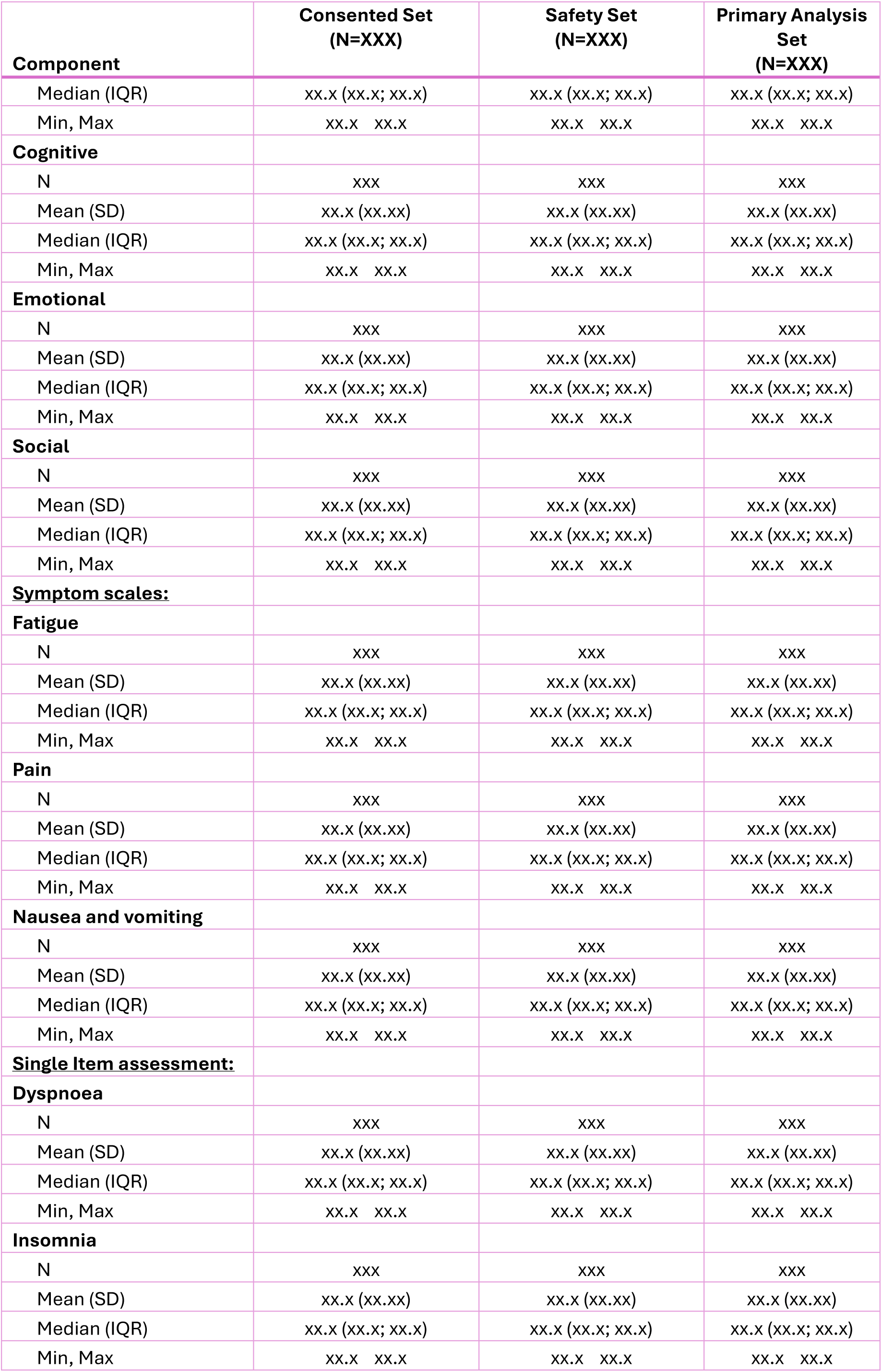
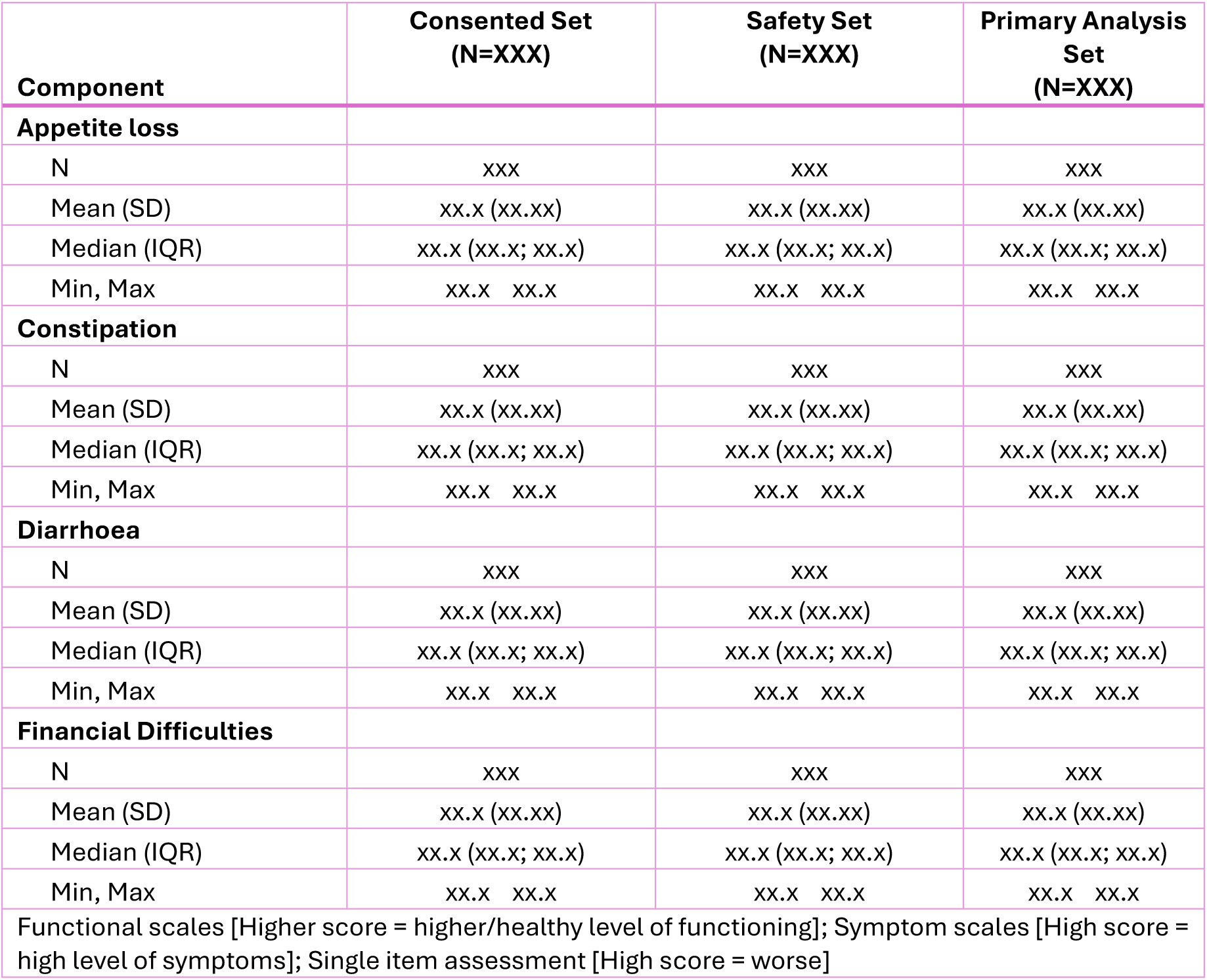
Baseline HRQOL: EORTC QLQ-C30 Summary.

| Component | Consented Set<br>(N=XXX) | Safety Set<br>(N=XXX) | Primary Analysis Set<br>(N=XXX) |
| --- | --- | --- | --- |
| **Global Health Status/QOL Scale** |  |  |  |
| N | xxx | xxx | xxx |
| Mean (SD) | xx.x (xx.xx) | xx.x (xx.xx) | xx.x (xx.xx) |
| Median (IQR) | xx.x (xx.x; xx.x) | xx.x (xx.x; xx.x) | xx.x (xx.x; xx.x) |
| Min, Max | xx.x xx.x | xx.x xx.x | xx.x xx.x |
| **QLQ-C30 Summary Score** |  |  |  |
| N | xxx | xxx | xxx |
| Mean (SD) | xx.x (xx.xx) | xx.x (xx.xx) | xx.x (xx.xx) |
| Median (IQR) | xx.x (xx.x; xx.x) | xx.x (xx.x; xx.x) | xx.x (xx.x; xx.x) |
| Min, Max | xx.x xx.x | xx.x xx.x | xx.x xx.x |
| **Functioning scales:** |  |  |  |
| **Physical** |  |  |  |
| N | xxx | xxx | xxx |
| Mean (SD) | xx.x (xx.xx) | xx.x (xx.xx) | xx.x (xx.xx) |
| Median (IQR) | xx.x (xx.x; xx.x) | xx.x (xx.x; xx.x) | xx.x (xx.x; xx.x) |
| Min, Max | xx.x xx.x | xx.x xx.x | xx.x xx.x |
| **Role** |  |  |  |
| N | xxx | xxx | xxx |
| Mean (SD) | xx.x (xx.xx) | xx.x (xx.xx) | xx.x (xx.xx) |

| **Component** | **Consented Set**<br>**(N=XXX)** | **Safety Set**<br>**(N=XXX)** | **Primary Analysis**<br>**Set**<br>**(N=XXX)** |
| --- | --- | --- | --- |
| Median (IQR) | xx.x (xx.x; xx.x) | xx.x (xx.x; xx.x) | xx.x (xx.x; xx.x) |
| Min, Max | xx.x xx.x | xx.x xx.x | xx.x xx.x |
| **Cognitive** |  |  |  |
| N | xxx | xxx | xxx |
| Mean (SD) | xx.x (xx.xx) | xx.x (xx.xx) | xx.x (xx.xx) |
| Median (IQR) | xx.x (xx.x; xx.x) | xx.x (xx.x; xx.x) | xx.x (xx.x; xx.x) |
| Min, Max | xx.x xx.x | xx.x xx.x | xx.x xx.x |
| **Emotional** |  |  |  |
| N | xxx | xxx | xxx |
| Mean (SD) | xx.x (xx.xx) | xx.x (xx.xx) | xx.x (xx.xx) |
| Median (IQR) | xx.x (xx.x; xx.x) | xx.x (xx.x; xx.x) | xx.x (xx.x; xx.x) |
| Min, Max | xx.x xx.x | xx.x xx.x | xx.x xx.x |
| **Social** |  |  |  |
| N | xxx | xxx | xxx |
| Mean (SD) | xx.x (xx.xx) | xx.x (xx.xx) | xx.x (xx.xx) |
| Median (IQR) | xx.x (xx.x; xx.x) | xx.x (xx.x; xx.x) | xx.x (xx.x; xx.x) |
| Min, Max | xx.x xx.x | xx.x xx.x | xx.x xx.x |
| **Symptom scales:** |  |  |  |
| **Fatigue** |  |  |  |
| N | xxx | xxx | xxx |
| Mean (SD) | xx.x (xx.xx) | xx.x (xx.xx) | xx.x (xx.xx) |
| Median (IQR) | xx.x (xx.x; xx.x) | xx.x (xx.x; xx.x) | xx.x (xx.x; xx.x) |
| Min, Max | xx.x xx.x | xx.x xx.x | xx.x xx.x |
| **Pain** |  |  |  |
| N | xxx | xxx | xxx |
| Mean (SD) | xx.x (xx.xx) | xx.x (xx.xx) | xx.x (xx.xx) |
| Median (IQR) | xx.x (xx.x; xx.x) | xx.x (xx.x; xx.x) | xx.x (xx.x; xx.x) |
| Min, Max | xx.x xx.x | xx.x xx.x | xx.x xx.x |
| **Nausea and vomiting** |  |  |  |
| N | xxx | xxx | xxx |
| Mean (SD) | xx.x (xx.xx) | xx.x (xx.xx) | xx.x (xx.xx) |
| Median (IQR) | xx.x (xx.x; xx.x) | xx.x (xx.x; xx.x) | xx.x (xx.x; xx.x) |
| Min, Max | xx.x xx.x | xx.x xx.x | xx.x xx.x |
| **Single Item assessment:** |  |  |  |
| **Dyspnoea** |  |  |  |
| N | xxx | xxx | xxx |
| Mean (SD) | xx.x (xx.xx) | xx.x (xx.xx) | xx.x (xx.xx) |
| Median (IQR) | xx.x (xx.x; xx.x) | xx.x (xx.x; xx.x) | xx.x (xx.x; xx.x) |
| Min, Max | xx.x xx.x | xx.x xx.x | xx.x xx.x |
| **Insomnia** |  |  |  |
| N | xxx | xxx | xxx |
| Mean (SD) | xx.x (xx.xx) | xx.x (xx.xx) | xx.x (xx.xx) |
| Median (IQR) | xx.x (xx.x; xx.x) | xx.x (xx.x; xx.x) | xx.x (xx.x; xx.x) |
| Min, Max | xx.x xx.x | xx.x xx.x | xx.x xx.x |
| **Appetite loss** |  |  |  |
| N | xxx | xxx | xxx |
| Mean (SD) | xx.x (xx.xx) | xx.x (xx.xx) | xx.x (xx.xx) |
| Median (IQR) | xx.x (xx.x; xx.x) | xx.x (xx.x; xx.x) | xx.x (xx.x; xx.x) |
| Min, Max | xx.x xx.x | xx.x xx.x | xx.x xx.x |
| **Constipation** |  |  |  |
| N | xxx | xxx | xxx |
| Mean (SD) | xx.x (xx.xx) | xx.x (xx.xx) | xx.x (xx.xx) |
| Median (IQR) | xx.x (xx.x; xx.x) | xx.x (xx.x; xx.x) | xx.x (xx.x; xx.x) |
| Min, Max | xx.x xx.x | xx.x xx.x | xx.x xx.x |
| **Diarrhoea** |  |  |  |
| N | xxx | xxx | xxx |
| Mean (SD) | xx.x (xx.xx) | xx.x (xx.xx) | xx.x (xx.xx) |
| Median (IQR) | xx.x (xx.x; xx.x) | xx.x (xx.x; xx.x) | xx.x (xx.x; xx.x) |
| Min, Max | xx.x xx.x | xx.x xx.x | xx.x xx.x |
| **Financial Difficulties** |  |  |  |
| N | xxx | xxx | xxx |
| Mean (SD) | xx.x (xx.xx) | xx.x (xx.xx) | xx.x (xx.xx) |
| Median (IQR) | xx.x (xx.x; xx.x) | xx.x (xx.x; xx.x) | xx.x (xx.x; xx.x) |
| Min, Max | xx.x xx.x | xx.x xx.x | xx.x xx.x |
| Functional scales [Higher score = higher/healthy level of functioning]; Symptom scales [High score = high level of symptoms]; Single item assessment [High score = worse] |  |  |  |

**Table 4.** Baseline HRQOL: EORTC QLQ PAN2c Summary.

| Component | Consented Set<br>(N=XXX) | Safety Set<br>(N=XXX) | Primary Analysis Set<br>(N=XXX) |
| --- | --- | --- | --- |
| **Multi-item scales:** |  |  |  |
| **Pancreatic pain** |  |  |  |
| N | xxx | xxx | xxx |
| Mean (SD) | xx.x (xx.xx) | xx.x (xx.xx) | xx.x (xx.xx) |
| Median (IQR) | xx.x (xx.x; xx.x) | xx.x (xx.x; xx.x) | xx.x (xx.x; xx.x) |
| Min, Max | xx.x xx.x | xx.x xx.x | xx.x xx.x |
| **Digestive symptoms** |  |  |  |
| N | xxx | xxx | xxx |
| Mean (SD) | xx.x (xx.xx) | xx.x (xx.xx) | xx.x (xx.xx) |
| Median (IQR) | xx.x (xx.x; xx.x) | xx.x (xx.x; xx.x) | xx.x (xx.x; xx.x) |
| Min, Max | xx.x xx.x | xx.x xx.x | xx.x xx.x |
| **Altered bowel habit** |  |  |  |
| N | xxx | xxx | xxx |
| Mean (SD) | xx.x (xx.xx) | xx.x (xx.xx) | xx.x (xx.xx) |
| Median (IQR) | xx.x (xx.x; xx.x) | xx.x (xx.x; xx.x) | xx.x (xx.x; xx.x) |
| Min, Max | xx.x xx.x | xx.x xx.x | xx.x xx.x |
| **Hepatic symptoms** |  |  |  |
| N | xxx | xxx | xxx |
| Mean (SD) | xx.x (xx.xx) | xx.x (xx.xx) | xx.x (xx.xx) |
| Median (IQR) | xx.x (xx.x; xx.x) | xx.x (xx.x; xx.x) | xx.x (xx.x; xx.x) |
| Min, Max | xx.x xx.x | xx.x xx.x | xx.x xx.x |
| **Body image** |  |  |  |
| N | xxx | xxx | xxx |
| Mean (SD) | xx.x (xx.xx) | xx.x (xx.xx) | xx.x (xx.xx) |
| Median (IQR) | xx.x (xx.x; xx.x) | xx.x (xx.x; xx.x) | xx.x (xx.x; xx.x) |
| Min, Max | xx.x xx.x | xx.x xx.x | xx.x xx.x |
| **Health care satisfaction** |  |  |  |
| N | xxx | xxx | xxx |
| Mean (SD) | xx.x (xx.xx) | xx.x (xx.xx) | xx.x (xx.xx) |
| Median (IQR) | xx.x (xx.x; xx.x) | xx.x (xx.x; xx.x) | xx.x (xx.x; xx.x) |
| Min, Max | xx.x xx.x | xx.x xx.x | xx.x xx.x |
| **Sexuality** |  |  |  |
| N | xxx | xxx | xxx |
| Mean (SD) | xx.x (xx.xx) | xx.x (xx.xx) | xx.x (xx.xx) |
| Median (IQR) | xx.x (xx.x; xx.x) | xx.x (xx.x; xx.x) | xx.x (xx.x; xx.x) |
| Min, Max | xx.x xx.x | xx.x xx.x | xx.x xx.x |
| **Single-item scales:** |  |  |  |
| **Flatulence** |  |  |  |
| N | xxx | xxx | xxx |
| Mean (SD) | xx.x (xx.xx) | xx.x (xx.xx) | xx.x (xx.xx) |
| Median (IQR) | xx.x (xx.x; xx.x) | xx.x (xx.x; xx.x) | xx.x (xx.x; xx.x) |
| Min, Max | xx.x xx.x | xx.x xx.x | xx.x xx.x |
| **Dry Mouth** |  |  |  |
| N | xxx | xxx | xxx |
| Mean (SD) | xx.x (xx.xx) | xx.x (xx.xx) | xx.x (xx.xx) |
| Median (IQR) | xx.x (xx.x; xx.x) | xx.x (xx.x; xx.x) | xx.x (xx.x; xx.x) |
| Min, Max | xx.x xx.x | xx.x xx.x | xx.x xx.x |
| **Indigestion** |  |  |  |
| N | xxx | xxx | xxx |
| Mean (SD) | xx.x (xx.xx) | xx.x (xx.xx) | xx.x (xx.xx) |
| Median (IQR) | xx.x (xx.x; xx.x) | xx.x (xx.x; xx.x) | xx.x (xx.x; xx.x) |
| Min, Max | xx.x xx.x | xx.x xx.x | xx.x xx.x |
| **Losing hair** |  |  |  |
| N | xxx | xxx | xxx |
| Mean (SD) | xx.x (xx.xx) | xx.x (xx.xx) | xx.x (xx.xx) |
| Median (IQR) | xx.x (xx.x; xx.x) | xx.x (xx.x; xx.x) | xx.x (xx.x; xx.x) |
| Min, Max | xx.x xx.x | xx.x xx.x | xx.x xx.x |
| **Weight loss** |  |  |  |
| N | xxx | xxx | xxx |
| Mean (SD) | xx.x (xx.xx) | xx.x (xx.xx) | xx.x (xx.xx) |
| Median (IQR) | xx.x (xx.x; xx.x) | xx.x (xx.x; xx.x) | xx.x (xx.x; xx.x) |
| Min, Max | xx.x xx.x | xx.x xx.x | xx.x xx.x |
| **Fear of future health** |  |  |  |
| N | xxx | xxx | xxx |
| Mean (SD) | xx.x (xx.xx) | xx.x (xx.xx) | xx.x (xx.xx) |
| Median (IQR) | xx.x (xx.x; xx.x) | xx.x (xx.x; xx.x) | xx.x (xx.x; xx.x) |
| Min, Max | xx.x xx.x | xx.x xx.x | xx.x xx.x |
| **Ability to plan future** |  |  |  |
| N | xxx | xxx | xxx |
| Mean (SD) | xx.x (xx.xx) | xx.x (xx.xx) | xx.x (xx.xx) |
| Median (IQR) | xx.x (xx.x; xx.x) | xx.x (xx.x; xx.x) | xx.x (xx.x; xx.x) |
| Min, Max | xx.x xx.x | xx.x xx.x | xx.x xx.x |
| Multi-item scales [High score = worse symptoms]; Single-item scales [High score = worse symptoms] |  |  |  |

**Table 5.** Baseline HRQOL: EQ5D-5L Summary by domain.

| Domain | Consented Set<br>(N=XX) | Safety Set<br>(N=XX) | Primary Analysis Set<br>(N=XX) |
| --- | --- | --- | --- |
| **Usual activities** |  |  |  |
| No problems | xxx/xxx (xx.x%) | xxx/xxx (xx.x%) | xxx/xxx (xx.x%) |
| Slight problems | xxx/xxx (xx.x%) | xxx/xxx (xx.x%) | xxx/xxx (xx.x%) |
| Moderate problems | xxx/xxx (xx.x%) | xxx/xxx (xx.x%) | xxx/xxx (xx.x%) |
| Severe problems | xxx/xxx (xx.x%) | xxx/xxx (xx.x%) | xxx/xxx (xx.x%) |
| Extreme problems | xxx/xxx (xx.x%) | xxx/xxx (xx.x%) | xxx/xxx (xx.x%) |
| **Overall Motility** |  |  |  |
| No problems | xxx/xxx (xx.x%) | xxx/xxx (xx.x%) | xxx/xxx (xx.x%) |
| Slight problems | xxx/xxx (xx.x%) | xxx/xxx (xx.x%) | xxx/xxx (xx.x%) |
| Moderate problems | xxx/xxx (xx.x%) | xxx/xxx (xx.x%) | xxx/xxx (xx.x%) |
| Severe problems | xxx/xxx (xx.x%) | xxx/xxx (xx.x%) | xxx/xxx (xx.x%) |
| Extreme problems | xxx/xxx (xx.x%) | xxx/xxx (xx.x%) | xxx/xxx (xx.x%) |
| **Pain/Discomfort** |  |  |  |
| No problems | xxx/xxx (xx.x%) | xxx/xxx (xx.x%) | xxx/xxx (xx.x%) |
| Slight problems | xxx/xxx (xx.x%) | xxx/xxx (xx.x%) | xxx/xxx (xx.x%) |
| Moderate problems | xxx/xxx (xx.x%) | xxx/xxx (xx.x%) | xxx/xxx (xx.x%) |
| Severe problems | xxx/xxx (xx.x%) | xxx/xxx (xx.x%) | xxx/xxx (xx.x%) |
| Extreme problems | xxx/xxx (xx.x%) | xxx/xxx (xx.x%) | xxx/xxx (xx.x%) |
| **Overall emotional health (Anxiety/Depression)** |  |  |  |
| No problems | xxx/xxx (xx.x%) | xxx/xxx (xx.x%) | xxx/xxx (xx.x%) |
| Slight problems | xxx/xxx (xx.x%) | xxx/xxx (xx.x%) | xxx/xxx (xx.x%) |
| Moderate problems | xxx/xxx (xx.x%) | xxx/xxx (xx.x%) | xxx/xxx (xx.x%) |
| Severe problems | xxx/xxx (xx.x%) | xxx/xxx (xx.x%) | xxx/xxx (xx.x%) |
| Extreme problems | xxx/xxx (xx.x%) | xxx/xxx (xx.x%) | xxx/xxx (xx.x%) |
| **Personal care** |  |  |  |
| No problems | xxx/xxx (xx.x%) | xxx/xxx (xx.x%) | xxx/xxx (xx.x%) |
| Slight problems | xxx/xxx (xx.x%) | xxx/xxx (xx.x%) | xxx/xxx (xx.x%) |
| Moderate problems | xxx/xxx (xx.x%) | xxx/xxx (xx.x%) | xxx/xxx (xx.x%) |
| Severe problems | xxx/xxx (xx.x%) | xxx/xxx (xx.x%) | xxx/xxx (xx.x%) |
| Extreme problems | xxx/xxx (xx.x%) | xxx/xxx (xx.x%) | xxx/xxx (xx.x%) |
| **Overall self-reported health score from EQ5D-5L (0 – 100)** |  |  |  |
| n | xxx | xxx | xxx |
| Mean (SD) | xx.x (xx.xx) | xx.x (xx.xx) | xx.x (xx.xx) |
| Median (Q1; Q3) | xx.x (xx.x; xx.x) | xx.x (xx.x; xx.x) | xx.x (xx.x; xx.x) |
| min max | xx.x xx.x | xx.x xx.x | xx.x xx.x |

**Table 6.** Patients’ previous anticancer therapy.

| Clinical characteristic | Consented Set<br>(N=XXX) | Safety Set<br>(N=XXX) | Primary Analysis Set<br>(N=XXX) |
| --- | --- | --- | --- |
| **Surgery** | xxx/xxx (xx.x%) | xxx/xxx (xx.x%) | xxx/xxx (xx.x%) |
| **Radiotherapy** | xxx/xxx (xx.x%) | xxx/xxx (xx.x%) | xxx/xxx (xx.x%) |
| **Indication** |  |  |  |
| Adjuvant | xxx/xxx (xx.x%) | xxx/xxx (xx.x%) | xxx/xxx (xx.x%) |
| Neoadjuvant | xxx/xxx (xx.x%) | xxx/xxx (xx.x%) | xxx/xxx (xx.x%) |
| Locally advanced disease | xxx/xxx (xx.x%) | xxx/xxx (xx.x%) | xxx/xxx (xx.x%) |
| Locally recurrent disease | xxx/xxx (xx.x%) | xxx/xxx (xx.x%) | xxx/xxx (xx.x%) |
| Metastatic disease | xxx/xxx (xx.x%) | xxx/xxx (xx.x%) | xxx/xxx (xx.x%) |
| Palliative | xxx/xxx (xx.x%) | xxx/xxx (xx.x%) | xxx/xxx (xx.x%) |
| **Reason for stopping** |  |  |  |
| Disease progression | xxx/xxx (xx.x%) | xxx/xxx (xx.x%) | xxx/xxx (xx.x%) |
| Regimen completed | xxx/xxx (xx.x%) | xxx/xxx (xx.x%) | xxx/xxx (xx.x%) |
| Toxicity | xxx/xxx (xx.x%) | xxx/xxx (xx.x%) | xxx/xxx (xx.x%) |
| Other | xxx/xxx (xx.x%) | xxx/xxx (xx.x%) | xxx/xxx (xx.x%) |
| **Systemic Therapy** | xxx/xxx (xx.x%) | xxx/xxx (xx.x%) | xxx/xxx (xx.x%) |
| **Indication** |  |  |  |
| Adjuvant | xxx/xxx (xx.x%) | xxx/xxx (xx.x%) | xxx/xxx (xx.x%) |
| Neoadjuvant | xxx/xxx (xx.x%) | xxx/xxx (xx.x%) | xxx/xxx (xx.x%) |
| Locally advanced disease | xxx/xxx (xx.x%) | xxx/xxx (xx.x%) | xxx/xxx (xx.x%) |
| Locally recurrent disease | xxx/xxx (xx.x%) | xxx/xxx (xx.x%) | xxx/xxx (xx.x%) |
| Metastatic disease | xxx/xxx (xx.x%) | xxx/xxx (xx.x%) | xxx/xxx (xx.x%) |
| Palliative | xxx/xxx (xx.x%) | xxx/xxx (xx.x%) | xxx/xxx (xx.x%) |
| **Reason for stopping** |  |  |  |
| Disease progression | xxx/xxx (xx.x%) | xxx/xxx (xx.x%) | xxx/xxx (xx.x%) |
| Regimen completed | xxx/xxx (xx.x%) | xxx/xxx (xx.x%) | xxx/xxx (xx.x%) |
| Toxicity | xxx/xxx (xx.x%) | xxx/xxx (xx.x%) | xxx/xxx (xx.x%) |
| Other | xxx/xxx (xx.x%) | xxx/xxx (xx.x%) | xxx/xxx (xx.x%) |
| **No. cycles completed** |  |  |  |
| N | xxx | xxx | xxx |
| Mean (SD) | xx.x (xx.xx) | xx.x (xx.xx) | xx.x (xx.xx) |
| Median (IQR) | xx.x (xx.x; xx.x) | xx.x (xx.x; xx.x) | xx.x (xx.x; xx.x) |
| Min, Max | xx.x xx.x | xx.x xx.x | xx.x xx.x |

**Table 7.** Summary of primary and secondary outcomes.

|  | Primary Analysis Set<br>(N=XXX) |
| --- | --- |
| **PFS 6months** | xxx/xxx (xx.x%) |
| **Objective tumour response rate** | xxx/xxx (xx.x%) |
| **Median PFS (months)** |  |
| N<sub>events</sub> | xxx |
| N<sub>censored</sub> | xxx |
| Median (95% CI) | xx.x (95% CI xx.x- xx.x) |
| **Overall Survival rate at 12months** | xxx/xxx (xx.x%) |
| **Median Overall Survival (months)** |  |
| N<sub>events</sub> | xxx |
| N<sub>censored</sub> | xxx |
| Median (95% CI) | xx.x (95% CI xx.x- xx.x) |
| **Time to progression (months)** |  |
| N<sub>events</sub> | xxx |
| N<sub>censored</sub> | xxx |
| Median (95% CI) | xx.x (95% CI xx.x- xx.x) |
| **Sulfasalazine dose intensity achieved (% of target dose)** |  |
| N | xxx |
| Mean (SD) | xx.x (xx.xx) |
| Median (IQR) | xx.x (xx.x; xx.x) |
| Min, Max | xx.x xx.x |

**Table 8.** Sulfasalazine dose intensity defined by acetylator status (secondary outcome)

|  | Slow Acetylator<br>(N=XXX) |  | Fast Acetylator<br>(N=XXX) |  | All Patients<br>(N=XXX) |  |
| --- | --- | --- | --- | --- | --- | --- |
|  | Dose intensity | Fraction of<br>intended dose (%) | Dose intensity | Fraction of<br>intended dose (%) | Dose intensity | Fraction of<br>intended dose (%) |
| **C1D4\*** |  |  |  |  |  |  |
| N | xxx | xxx | xxx | xxx | xxx | xxx |
| Mean (SD) | xx.x (xx.xx) | xx.x (xx.xx) | xx.x (xx.xx) | xx.x (xx.xx) | xx.x (xx.xx) | xx.x (xx.xx) |
| Median (IQR) | xx.x (xx.x; xx.x) | xx.x (xx.x; xx.x) | xx.x (xx.x; xx.x) | xx.x (xx.x; xx.x) | xx.x (xx.x; xx.x) | xx.x (xx.x; xx.x) |
| Min, Max | xx.x xx.x | xx.x xx.x | xx.x xx.x | xx.x xx.x | xx.x xx.x | xx.x xx.x |
| **C1D8** |  |  |  |  |  |  |
| N | xxx | xxx | xxx | xxx | xxx | xxx |
| Mean (SD) | xx.x (xx.xx) | xx.x (xx.xx) | xx.x (xx.xx) | xx.x (xx.xx) | xx.x (xx.xx) | xx.x (xx.xx) |
| Median (IQR) | xx.x (xx.x; xx.x) | xx.x (xx.x; xx.x) | xx.x (xx.x; xx.x) | xx.x (xx.x; xx.x) | xx.x (xx.x; xx.x) | xx.x (xx.x; xx.x) |
| Min, Max | xx.x xx.x | xx.x xx.x | xx.x xx.x | xx.x xx.x | xx.x xx.x | xx.x xx.x |
| **C1D11\*** |  |  |  |  |  |  |
| N | xxx | xxx | xxx | xxx | xxx | xxx |
| Mean (SD) | xx.x (xx.xx) | xx.x (xx.xx) | xx.x (xx.xx) | xx.x (xx.xx) | xx.x (xx.xx) | xx.x (xx.xx) |
| Median (IQR) | xx.x (xx.x; xx.x) | xx.x (xx.x; xx.x) | xx.x (xx.x; xx.x) | xx.x (xx.x; xx.x) | xx.x (xx.x; xx.x) | xx.x (xx.x; xx.x) |
| Min, Max | xx.x xx.x | xx.x xx.x | xx.x xx.x | xx.x xx.x | xx.x xx.x | xx.x xx.x |
| **C1D15** |  |  |  |  |  |  |
| N | xxx | xxx | xxx | xxx | xxx | xxx |
| Mean (SD) | xx.x (xx.xx) | xx.x (xx.xx) | xx.x (xx.xx) | xx.x (xx.xx) | xx.x (xx.xx) | xx.x (xx.xx) |
| Median (IQR) | xx.x (xx.x; xx.x) | xx.x (xx.x; xx.x) | xx.x (xx.x; xx.x) | xx.x (xx.x; xx.x) | xx.x (xx.x; xx.x) | xx.x (xx.x; xx.x) |
| Min, Max | xx.x xx.x | xx.x xx.x | xx.x xx.x | xx.x xx.x | xx.x xx.x | xx.x xx.x |
| **C1D18\*** |  |  |  |  |  |  |
| N | xxx | xxx | xxx | xxx | xxx | xxx |
| Mean (SD) | xx.x (xx.xx) | xx.x (xx.xx) | xx.x (xx.xx) | xx.x (xx.xx) | xx.x (xx.xx) | xx.x (xx.xx) |
| Median (IQR) | xx.x (xx.x; xx.x) | xx.x (xx.x; xx.x) | xx.x (xx.x; xx.x) | xx.x (xx.x; xx.x) | xx.x (xx.x; xx.x) | xx.x (xx.x; xx.x) |
| Min, Max | xx.x xx.x | xx.x xx.x | xx.x xx.x | xx.x xx.x | xx.x xx.x | xx.x xx.x |
| **C1D22** |  |  |  |  |  |  |
| N | xxx | xxx | xxx | xxx | xxx | xxx |
| Mean (SD) | xx.x (xx.xx) | xx.x (xx.xx) | xx.x (xx.xx) | xx.x (xx.xx) | xx.x (xx.xx) | xx.x (xx.xx) |
| Median (IQR) | xx.x (xx.x; xx.x) | xx.x (xx.x; xx.x) | xx.x (xx.x; xx.x) | xx.x (xx.x; xx.x) | xx.x (xx.x; xx.x) | xx.x (xx.x; xx.x) |
| Min, Max | xx.x xx.x | xx.x xx.x | xx.x xx.x | xx.x xx.x | xx.x xx.x | xx.x xx.x |
| **C2D1^** |  |  |  |  |  |  |
| N | xxx | xxx | xxx | xxx | xxx | xxx |
| Mean (SD) | xx.x (xx.xx) | xx.x (xx.xx) | xx.x (xx.xx) | xx.x (xx.xx) | xx.x (xx.xx) | xx.x (xx.xx) |
| Median (IQR) | xx.x (xx.x; xx.x) | xx.x (xx.x; xx.x) | xx.x (xx.x; xx.x) | xx.x (xx.x; xx.x) | xx.x (xx.x; xx.x) | xx.x (xx.x; xx.x) |
| Min, Max | xx.x xx.x | xx.x xx.x | xx.x xx.x | xx.x xx.x | xx.x xx.x | xx.x xx.x |
| **C2D15^** |  |  |  |  |  |  |
| N | xxx | xxx | xxx | xxx | xxx | xxx |
| Mean (SD) | xx.x (xx.xx) | xx.x (xx.xx) | xx.x (xx.xx) | xx.x (xx.xx) | xx.x (xx.xx) | xx.x (xx.xx) |
| Median (IQR) | xx.x (xx.x; xx.x) | xx.x (xx.x; xx.x) | xx.x (xx.x; xx.x) | xx.x (xx.x; xx.x) | xx.x (xx.x; xx.x) | xx.x (xx.x; xx.x) |
| Min, Max | xx.x xx.x | xx.x xx.x | xx.x xx.x | xx.x xx.x | xx.x xx.x | xx.x xx.x |
| **C3D1^** |  |  |  |  |  |  |
| N | xxx | xxx | xxx | xxx | xxx | xxx |
| Mean (SD) | xx.x (xx.xx) | xx.x (xx.xx) | xx.x (xx.xx) | xx.x (xx.xx) | xx.x (xx.xx) | xx.x (xx.xx) |
| Median (IQR) | xx.x (xx.x; xx.x) | xx.x (xx.x; xx.x) | xx.x (xx.x; xx.x) | xx.x (xx.x; xx.x) | xx.x (xx.x; xx.x) | xx.x (xx.x; xx.x) |
| Min, Max | xx.x xx.x | xx.x xx.x | xx.x xx.x | xx.x xx.x | xx.x xx.x | xx.x xx.x |
| **C4D1^** |  |  |  |  |  |  |
| N | xxx | xxx | xxx | xxx | xxx | xxx |
| Mean (SD) | xx.x (xx.xx) | xx.x (xx.xx) | xx.x (xx.xx) | xx.x (xx.xx) | xx.x (xx.xx) | xx.x (xx.xx) |
| Median (IQR) | xx.x (xx.x; xx.x) | xx.x (xx.x; xx.x) | xx.x (xx.x; xx.x) | xx.x (xx.x; xx.x) | xx.x (xx.x; xx.x) | xx.x (xx.x; xx.x) |
| Min, Max | xx.x xx.x | xx.x xx.x | xx.x xx.x | xx.x xx.x | xx.x xx.x | xx.x xx.x |
| **C5D1^** |  |  |  |  |  |  |
| N | xxx | xxx | xxx | xxx | xxx | xxx |
| Mean (SD) | xx.x (xx.xx) | xx.x (xx.xx) | xx.x (xx.xx) | xx.x (xx.xx) | xx.x (xx.xx) | xx.x (xx.xx) |
| Median (IQR) | xx.x (xx.x; xx.x) | xx.x (xx.x; xx.x) | xx.x (xx.x; xx.x) | xx.x (xx.x; xx.x) | xx.x (xx.x; xx.x) | xx.x (xx.x; xx.x) |
| Min, Max | xx.x xx.x | xx.x xx.x | xx.x xx.x | xx.x xx.x | xx.x xx.x | xx.x xx.x |

| C6D1^ |  |  |  |  |  |  |
| --- | --- | --- | --- | --- | --- | --- |
| N | xxx | xxx | xxx | xxx | xxx | xxx |
| Mean (SD) | xx.x (xx.xx) | xx.x (xx.xx) | xx.x (xx.xx) | xx.x (xx.xx) | xx.x (xx.xx) | xx.x (xx.xx) |
| Median (IQR) | xx.x (xx.x; xx.x) | xx.x (xx.x; xx.x) | xx.x (xx.x; xx.x) | xx.x (xx.x; xx.x) | xx.x (xx.x; xx.x) | xx.x (xx.x; xx.x) |
| Min, Max | xx.x xx.x | xx.x xx.x | xx.x xx.x | xx.x xx.x | xx.x xx.x | xx.x xx.x |
| ^Dose intensity is defined as absolute drug dose delivered per time unit expressed as g per week); **Primary Analysis Set** used for analysis |  |  |  |  |  |  |

**Table 9.** Summary of peripheral blood Glutathione levels.

| **Timepoint** | **Primary Analysis Set**<br>**(N=XXX)** |
| --- | --- |
| Pre-sulfasalazine |  |
| N | xxx |
| Mean (SD) | xx.x (xx.xx) |
| Median (IQR) | xx.x (xx.x; xx.x) |
| Min, Max | xx.x xx.x |
| Post-sulfasalazine (C2) |  |
| N | xxx |
| Mean (SD) | xx.x (xx.xx) |
| Median (IQR) | xx.x (xx.x; xx.x) |
| Min, Max | xx.x xx.x |
| Post-sulfasalazine (C3D1) |  |
| N | xxx |
| Mean (SD) | xx.x (xx.xx) |
| Median (IQR) | xx.x (xx.x; xx.x) |
| Min, Max | xx.x xx.x |
| Pre-post change: Post-sulfasalazine (C2) |  |
| N | xxx |
| Mean (SD) | xx.x (xx.xx) |
| Median (IQR) | xx.x (xx.x; xx.x) |
| Min, Max | xx.x xx.x |
| Pre-post change: Post-sulfasalazine (C3D1) |  |
| N | xxx |
| Mean (SD) | xx.x (xx.xx) |
| Median (IQR) | xx.x (xx.x; xx.x) |
| Min, Max | xx.x xx.x |

**Table 10.** Summary of serum collagen degradation levels.

| Timepoint | Type I<br>(N=XXX) | Type III<br>(N=XXX) | Type IV (C4M, C4M12a1)<br>(N=XXX) |
| --- | --- | --- | --- |
| Pre-sulfasalazine |  |  |  |
| N | xxx | xxx | xxx |
| Mean (SD) | xx.x (xx.xx) | xx.x (xx.xx) | xx.x (xx.xx) |
| Median (IQR) | xx.x (xx.x; xx.x) | xx.x (xx.x; xx.x) | xx.x (xx.x; xx.x) |
| Min, Max | xx.x xx.x | xx.x xx.x | xx.x xx.x |
| Post-sulfasalazine (C3D1/PDV) |  |  |  |
| N | xxx | xxx | xxx |
| Mean (SD) | xx.x (xx.xx) | xx.x (xx.xx) | xx.x (xx.xx) |
| Median (IQR) | xx.x (xx.x; xx.x) | xx.x (xx.x; xx.x) | xx.x (xx.x; xx.x) |
| Min, Max | xx.x xx.x | xx.x xx.x | xx.x xx.x |
| <sup>1</sup>Primary Analysis Set used for analysis |  |  |  |

**Table 11.** Summary of Tumour marker CA 1S-S and CEA.

| Clinical characteristic | CA 19-9<br>(N=XXX) | CEA<br>(N=XXX) |
| --- | --- | --- |
| >2x upper limit of normal (ULN) at screening | xxx/xxx (xx.x%) | xxx/xxx (xx.x%) |
| **Reduction from baseline to nadir ≥:** |  |  |
| 0% | xxx/xxx (xx.x%) | xxx/xxx (xx.x%) |
| 20% | xxx/xxx (xx.x%) | xxx/xxx (xx.x%) |
| 60% | xxx/xxx (xx.x%) | xxx/xxx (xx.x%) |
| 90% | xxx/xxx (xx.x%) | xxx/xxx (xx.x%) |
| **Tumour marker level** |  |  |
| **Baseline** |  |  |
| N | xxx | xxx |
| Mean (SD) | xx.x (xx.xx) | xx.x (xx.xx) |
| Median (IQR) | xx.x (xx.x; xx.x) | xx.x (xx.x; xx.x) |
| Min, Max | xx.x xx.x | xx.x xx.x |
| **C1D1** |  |  |
| N | xxx | xxx |
| Mean (SD) | xx.x (xx.xx) | xx.x (xx.xx) |
| Median (IQR) | xx.x (xx.x; xx.x) | xx.x (xx.x; xx.x) |
| Min, Max | xx.x xx.x | xx.x xx.x |
| **C2D1** |  |  |
| N | xxx | xxx |
| Mean (SD) | xx.x (xx.xx) | xx.x (xx.xx) |
| Median (IQR) | xx.x (xx.x; xx.x) | xx.x (xx.x; xx.x) |
| Min, Max | xx.x xx.x | xx.x xx.x |
| **C3D1** |  |  |
| N | xxx | xxx |
| Mean (SD) | xx.x (xx.xx) | xx.x (xx.xx) |
| Median (IQR) | xx.x (xx.x; xx.x) | xx.x (xx.x; xx.x) |
| Min, Max | xx.x xx.x | xx.x xx.x |
| **C4D1** |  |  |
| N | xxx | xxx |
| Mean (SD) | xx.x (xx.xx) | xx.x (xx.xx) |
| Median (IQR) | xx.x (xx.x; xx.x) | xx.x (xx.x; xx.x) |
| Min, Max | xx.x xx.x | xx.x xx.x |
| **C5D1** |  |  |
| N | xxx | xxx |
| Mean (SD) | xx.x (xx.xx) | xx.x (xx.xx) |
| Median (IQR) | xx.x (xx.x; xx.x) | xx.x (xx.x; xx.x) |
| Min, Max | xx.x xx.x | xx.x xx.x |
| **C6D1** |  |  |
| N | xxx | xxx |
| Mean (SD) | xx.x (xx.xx) | xx.x (xx.xx) |
| Median (IQR) | xx.x (xx.x; xx.x) | xx.x (xx.x; xx.x) |
| Min, Max | xx.x xx.x | xx.x xx.x |
| <sup>1</sup>Primary Analysis Set used for analysis |  |  |

**Table 12.** Summary of time on study by line of therapy.

| Line of therapy | N | Mean (SD) | Median (IQR) |
| --- | --- | --- | --- |
| Second-line | x | xx.x (xx.xx) | xx.x (xx.x; xx.x) |
| Beyond second-line (ECOG 0-1) | x | xx.x (xx.xx) | xx.x (xx.x; xx.x) |
| <sup>1</sup>Consented Set used for analysis |  |  |  |

**Table 13.** Post hoc linear regression analysis of time on study by baseline characteristics.

| Baseline Characteristic | Estimate ($\beta$) 95% CI | p-value |
| --- | --- | --- |
| Line of Therapy | xx.x (xx - xx) | x.xxx |
| ECOG Performance status (0-1 vs. 2-3) | xx.x (xx - xx) | x.xxx |
| Albumin | xx.x (xx - xx) | x.xxx |
| White cell count | xx.x (xx - xx) | x.xxx |
| QOL-C30 summary score | xx.x (xx - xx) | x.xxx |
| <sup>1</sup>Consented Set used for analysis; CI= confidence interval |  |  |

**Table 14.** Post hoc Kaplan-Meier survival analysis of overall survival by baseline characteristics.

| Baseline Characteristic | N | N Events | N Censored | Median Survival, Months (95% CI) | Overall Comparison<sup>2</sup> |  |
| --- | --- | --- | --- | --- | --- | --- |
|  |  |  |  |  | Chi-square | p-value |
| **SLC7A11** |  |  |  |  | xx.x | x.xxx |
| Group 1 | xx | xx | xx | xx.x (xx - xx) |  |  |
| Group 2 | xx | xx | xx | xx.x (xx - xx) |  |  |
| **Tumour metabolic response** |  |  |  |  | xx.x | x.xxx |
| Group 1 | xx | xx | xx | xx.x (xx - xx) |  |  |
| Group 2 | xx | xx | xx | xx.x (xx - xx) |  |  |
| **Serum collagen degradation** |  |  |  |  |  |  |
| **Type I** |  |  |  |  | xx.x | x.xxx |
| Group 1 | xx | xx | xx | xx.x (xx - xx) |  |  |
| Group 2 | xx | xx | xx | xx.x (xx - xx) |  |  |
| **Type III** |  |  |  |  | xx.x | x.xxx |
| Group 1 | xx | xx | xx | xx.x (xx - xx) |  |  |
| Group 2 | xx | xx | xx | xx.x (xx - xx) |  |  |
| **Type IV (C4M, C4M12a1)** |  |  |  |  | xx.x | x.xxx |
| Group 1 | xx | xx | xx | xx.x (xx - xx) |  |  |
| Group 2 | xx | xx | xx | xx.x (xx - xx) |  |  |
| <sup>1</sup>Consented Set used for analysis; <sup>2</sup>Log-rank test or alternate weighted log-rank tests; CI= confidence interval |  |  |  |  |  |  |

**Table 15.** Post hoc Logistic regression analysis of remaining on therapy for at least one month by baseline characteristics.

| Baseline Characteristic | N | OR (95% CI) | AUC (95% CI) | p-value |
| --- | --- | --- | --- | --- |
| Platelet-to-lymphocyte ratio (POR) | xx | xx.x (xx - xx) | xx.x (xx - xx) | x.xxx |
| White cell count (WCC) | xx | xx.x (xx - xx) | xx.x (xx - xx) | x.xxx |
| Serum albumin | xx | xx.x (xx - xx) | xx.x (xx - xx) | x.xxx |
| Liver function test (LFT) | xx | xx.x (xx - xx) | xx.x (xx - xx) | x.xxx |
| <sup>1</sup>Safety Set used for analysis; OR= Odds ratio, CI= confidence interval, AUC = area under curve |  |  |  |  |

**Table 16.** Adverse events.

|  | Safety Set<br>(N=XXX) |
| --- | --- |
| **Any Serious Adverse Event (SAE)** |  |
| Yes | xx# xxx/xxx (xx.x%) |
| No | xx# xxx/xxx (xx.x%) |
| **Seriousness criteria** |  |
| Death not related to PDAC | xx# xxx/xxx (xx.x%) |
| Life-threatening | xx# xxx/xxx (xx.x%) |
| Hospitalisation or prolongation of hospitalisation | xx# xxx/xxx (xx.x%) |
| Significant disability or incapacity | xx# xxx/xxx (xx.x%) |
| Congenital Abnormality | xx# xxx/xxx (xx.x%) |
| Other Important Medical Event | xx# xxx/xxx (xx.x%) |
| **Causal Relationship to study medication** |  |
| Definitely related | xx# xxx/xxx (xx.x%) |
| Probably related | xx# xxx/xxx (xx.x%) |
| Possibly related | xx# xxx/xxx (xx.x%) |
| Unlikely to be related | xx# xxx/xxx (xx.x%) |
| Definitely not related | xx# xxx/xxx (xx.x%) |
| **AE Severity (NCI CTCAE v5.0)** |  |
| Grade 1 – Mild | xx# xxx/xxx (xx.x%) |
| Grade 2 – Moderate | xx# xxx/xxx (xx.x%) |
| Grade 3 – Severe or medically significant | xx# xxx/xxx (xx.x%) |
| Grade 4 – Life-threatening consequences | xx# xxx/xxx (xx.x%) |
| Grade 5 – Death related to AE | xx# xxx/xxx (xx.x%) |
| **AE expected per sulfasalazine product information?** |  |
| No | xx# xxx/xxx (xx.x%) |
| Yes | xx# xxx/xxx (xx.x%) |
| **Event status** |  |
| Ongoing | xx# xxx/xxx (xx.x%) |
| Resolved | xx# xxx/xxx (xx.x%) |
| Resolved with sequelae | xx# xxx/xxx (xx.x%) |
| Death | xx# xxx/xxx (xx.x%) |
| Unsure | xx# xxx/xxx (xx.x%) |
| **Action taken with sulfasalazine** |  |
| Temporarily stopped | xx# xxx/xxx (xx.x%) |

|  | **Safety Set**<br>**(N=XXX)** |
| --- | --- |
| Dose reduced | xx# xxx/xxx (xx.x%) |
| Permanently discontinued | xx# xxx/xxx (xx.x%) |
| No action taken | xx# xxx/xxx (xx.x%) |
| **Dose down-titrated to** |  |
| 4.5g | xx# xxx/xxx (xx.x%) |
| 3.0g | xx# xxx/xxx (xx.x%) |
| 1.5g | xx# xxx/xxx (xx.x%) |
| Other | xx# xxx/xxx (xx.x%) |
| **If permanently discontinued, temporarily stopped or dose reduced, did the reaction abate?** |  |
| No | xx# xxx/xxx (xx.x%) |
| Yes | xx# xxx/xxx (xx.x%) |
| **If temporarily stopped or dose reduced, did the reaction re-appear after re-introduction?** |  |
| No | xx# xxx/xxx (xx.x%) |
| Yes | xx# xxx/xxx (xx.x%) |
| Notes: All AEs are summarised as the total number of events reported# followed by the number and percentage of patients experiencing at least one event. The denominators for percentages are all randomised patients. |  |

---

**Listing 1:** Adverse events
| Patient ID | Diagnosis | SAE | Seriousness criteria | AE Start Date | Date serious criteria met | Date admitted to hospital | Date of Discharge | Date of Death | Severity | Event status | Causality | Action taken with study drug | Dose at time of event | Dose down-titrated to | Did reaction abate? | Did reaction re-appear after re-introduction? | Event narrative |
| --- | --- | --- | --- | --- | --- | --- | --- | --- | --- | --- | --- | --- | --- | --- | --- | --- | --- |
| <sup>1</sup> Safety Set population displayed |  |  |  |  |  |  |  |  |  |  |  |  |  |  |  |  |  |

| Patient ID | ECOG performance status | POR | WCC | Albumin | Liver function test | Date of Screening | Date of Enrolment | Date of Study Drug Administration | Date of Study Discontinuation | Follow-up time |
| --- | --- | --- | --- | --- | --- | --- | --- | --- | --- | --- |
| <sup>1</sup> Screening Set population displayed |  |  |  |  |  |  |  |  |  |  |

